# Monocyte and neutrophil levels are potentially linked to progression to IPF for patients with indeterminate UIP CT pattern

**DOI:** 10.1101/2021.08.20.21262390

**Authors:** A. Achaiah, A. Rathnapala, A. Pereira, H. Bothwell, K. Dwivedi, R. Barker, R Benamore, R. Hoyles, V Iotchkova, L.P. Ho

**Affiliations:** MRC Human Immunology Unit, Weatherall Institute of Molecular Medicine, University of Oxford; Oxford Interstitial Lung Disease Service, Oxford University Hospitals NHS Foundation Trust, Oxford; Thoracic Radiology Department, Oxford University Hospitals NHS Foundation Trust, Oxford; Computational Biology and Statistics Unit, Weatherall Institute of Molecular Medicine, University of Oxford

**Keywords:** Interstitial Fibrosis, Innate Immunity

## Abstract

**Rationale:** Idiopathic pulmonary fibrosis (IPF) is a progressive fibrotic lung disease with poor prognosis. Identifying patients early may allow intervention which could limit progression. The ‘indeterminate for UIP’ (iUIP) CT pattern, defined in the 2018 IPF guidelines, could be a precursor to IPF but there is limited data on how patients with iUIP progress over time.

**Objective:** To evaluate the radiological progression of iUIP and explore factors linked to progression to IPF.

**Methods:** We performed a retrospective analysis of a lung fibrosis clinic cohort (n=230) seen between 2013-2017. Cases with iUIP were identified; first ever CTs for each patient found and categorised as ‘non-progressor’ or ‘progressors’ (the latter defined as increase in extent of disease or to ‘definite’ or ‘probable’ UIP CT pattern) during their follow up. Lung function trends, haematological data and patient demographics were examined to explore disease evolution and potential contribution to progression.

**Results:** 48 cases with iUIP CT pattern were identified. Of these, 32 had follow up CT scans, of which 23 demonstrated progression. 17 patients in this cohort were diagnosed with IPF over a mean (S.D.) period of 3.9 (±1.9) years. Monocyte [HR 23, CI 1.6-340, p=0.03] and neutrophil levels [HR 1.8, CI 1.3-2.3, p<0.001] obtained around the time of initial CT, were associated with progression to IPF using Cox proportional hazard modelling.

**Conclusion:** 53% of our evaluable iUIP patients progressed to IPF over a mean of four years. Monocyte and neutrophil levels at initial CT were significantly associated with progression in disease. These data provide a single-centre analysis of the evolution of patients with iUIP CT pattern, and first signal for potential factors associated with progression to IPF.

**Key message:** *What is the key question?:* How does the ‘indeterminate for UIP’ (iUIP) interstitial CT pattern evolve over time and what factors are associated with progression to definite and probable UIP pattern.

*What is the bottom line?:* In this retrospective single centre analysis, 53% of evaluable cases with iUIP on initial CT scan progressed to probable or definite UIP CT pattern over an average of 4 years. Monocyte and neutrophil levels performed around the time of initial CT were significantly associated with progression to definite and probable UIP pattern.

*Why read on?:* We discuss the implications of these findings, its strengths and limitations.

## Background

Idiopathic pulmonary fibrosis (IPF) is a progressive fibrotic condition characterised by a distinctive fibrotic pattern on thoracic CT scans, referred to as ‘Usual Interstitial Pneumonia’ (UIP). Despite advances in treatment, prognosis remains poor with a median survival of 2-4 years from diagnosis^1^. Identifying and treating patients earlier could improve outcome.

It has been long acknowledged that there is a group of patients with subclinical interstitial lung disease. The term ‘interstitial lung abnormalities’ (ILA) was originally coined to define the spectrum of radiological patterns seen in these patients^2^. ‘Indeterminate for UIP’ (iUIP) is one of these ILA subtypes. iUIP CT pattern is defined by presence of subtle reticulation, in the absence of honeycombing and traction bronchiectasis, with or without mild ground glass opacification (GGO) in a basal and sub-pleural distribution. The term was included in the 2018 ATS/ERS/JRS/ALAT IPF guideline to categorise CT features that do not meet the criteria for ‘definite’ or ‘probable’ UIP, and where there is no alternative ILD diagnosis^3^. Prior to the 2018 guideline, cases compatible with the current iUIP and ‘probable UIP’ definition were collectively categorised as ‘possible UIP’^4^. This was a highly heterogenous group, and many cases were subjected to surgical lung biopsies^5^ for clarification of diagnosis. Data accumulated from these biopsies suggest that in a significant number of patients with possible UIP pattern and particularly those with traction bronchiectasis, there is a reasonably good association with a histology diagnosis of UIP^6,7^. As a result, the 2018 guidelines specifies that where there is traction bronchiectasis, ‘possible UIP’ CT patterns should be placed into the category of ‘probable UIP’. This leaves those with no traction bronchiectasis as ‘iUIP’ with features described above. Little is known of how radiographic iUIP progresses, although a study showed that up to 30% of cases that were biopsied demonstrated a histology pattern of UIP^6^. This indicates that at least a proportion of patients with iUIP progresses to a clinical diagnosis of IPF. It is not clear which factors are associated with this^8^.

In this retrospective cohort study, we evaluated the radiological and clinical progression of patients with the iUIP CT pattern in one Interstitial Lung Disease centre, dividing cases into ‘non-progressors’ and ‘progressors’ (to ‘probable’ and ‘definite’ UIP pattern on CT by 2018 criteria or in extent of fibrosis on CT). We explored the association between blood neutrophil, monocytes and lymphocyte levels near to the point of first CT and patient demographics with progression. We found that 53% of evaluable cases progressed to a CT pattern of ‘probable’ or ‘definite UIP’ (all with IPF clinical diagnosis) within four years (mean/S.D of 3.9/1.5 years) of initial CT. Using Cox proportional hazard analysis, we found that neutrophil and monocytes (but not lymphocytes), measured within 3 months of initial CT, significantly correlated with progression of iUIP in extent, and to a diagnosis of IPF.

## Methods

### Study design

We first, identified patients with iUIP CT patterns amongst our lung fibrosis cohort of patients who attended the Oxford Interstitial Lung Disease Service between 2013 and 2017. All patients in this lung fibrosis cohort had a CT pattern of ‘possible UIP’ or ‘UIP’ according to 2011. Radiologist reports for all available thoracic CTs for these patients (including first CTs before 2013) and up to August 2019 were re-analysed and cross-checked with reports from ILD multidisciplinary team meetings (MDT), and re-categorised according to the 2018 IPF guideline^3^. Patients with iUIP CT patterns were then grouped as either ‘non-progressive’ or ‘progressive’ based on comparison of their first CT (including those prior to attendance at Oxford) to the latest follow-on CT (up to cut off point of August 2019). We defined ‘non-progressive’ as no change in CT scan in terms of extent of disease or change in pattern of disease; and ‘progressive’ if there were either visual (qualitative) increase in extent of disease or progression of CT pattern to ‘definite’ or ‘probable UIP’ pattern.

The following data were collected - patient demographics, dates of first CT and follow-on CTs, year of diagnosis of IPF (by ILD MDT, and according to 2018 guidelines), all available pulmonary function tests (PFTs), co-morbidities, neutrophils, lymphocytes and monocyte levels performed using standard hospital ‘full blood count’ analysis and clinical outcomes including disease progression, survival and years of clinic follow up. Indications for follow on CT were also recorded.

### Patient and Public Involvement

Patient and public were not involved in design, recruitment, conduct of this study.

### Ethical approval

The study was part of a study to examine the factors associated with disease progression in IPF (ethical approval 14/SC/1060 from the Health Research Authority and South-Central National Research Ethics Service).

### CT scan and analysis

CT scans were acquired using a 64-detector row CT scanner (LightSpeed VCT XT; GE Medical Systems, Milwaukee, WI, USA). Images were reconstructed using a high spatial resolution algorithm. All CT abnormalities were defined and analysed using standard Fleischner-based terminology^9^.

### Statistical analysis

Where data are expressed as means, standard deviation are shown. Tests for normality of data was performed using a D’Agostino & Pearson test and following this the difference between groups was analysed using Student’s t-tests or Mann-Whitney test for parametric and non-parametric analysis respectively. Contingency tests (Fisher’s exact test of significance) were used to assess categorical data. Survival analysis (Log rank test of significance) was performed to evaluate time-to-mortality. For analyses of correlates for progression to IPF (2018 criteria)^2^, the Cox proportional hazard modelling to determine hazard ratios (HRs) was employed, testing their significance in two settings: (A) using data from patients regardless of whether they proceeded to defined events (‘definite’ or ‘probable’ UIP pattern on CT or progression in extent of disease) but up to August 2019 (n=32) and (B) restricting the patients to only those that did progress to ‘definite’ or ‘probable’ UIP pattern on CT during the period of analysis (n=17). Hazard ratios generated for continuous covariates represent the change in the risk of outcome if the covariate in question changes by one unit. Hazard ratios generated for dichotomised co-variates represents the risk of achieving outcome if the co-variate is present. All analyses were performed using Graphpad Prism (version 8) with the exception of Cox proportional hazard modelling which was performed with R Studio (version 3.6.2) statistical programming language packages Survival (3.1.8) and Survminer (0.4.6) by our statistical team (led by VI). We used the cox.zph function in the R package survival for testing the proportional hazards assumption of a Cox regression. Statistical significance was performed using the likelihood ratio test (preferred compared to Wald test due to our smaller number) as reported by the coxph function at the 95% significance level. Reported p values were two-sided and a p value <0.05 was considered significant.

## Results

### Progress of iUIP patients from point of initial CTs

Of the 230 individual patients who attended the lung fibrosis ILD clinic between 2013 and 2017 with CT patterns of ‘probable’, ‘possible’ or ‘definite UIP’, 48 (21%) cases with iUIP pattern were found. 32 of these patients had at least one follow-on thoracic CT from the first scan. In the 16 cases that did not have a follow-on scan (and therefore could not be categorised into progressors or non-progressors), 13 cases were discharged after a mean of 2.1 years without a CT, as they were clinically stable and three were deemed not to require a second CT scan clinically.

Of the 32 patients that had follow-on scans, 9 (28%) patients showed no change in CT pattern or extent in disease over a mean (±S.D.) of 2.1 ±0.9 years and were classified as non-progressors. The most frequent indications for follow-on CT in this group was for nodule surveillance (64%) and to identify any further radiographic progression of iUIP (14%). 23 (72%) demonstrated progression. 6 showed increase in extent of iUIP but no change in pattern over 3.1 ±0.8 yrs; 11 progressed to ‘probable UIP’ over 3.8 ±1.6 years and 6 to ‘definite’ UIP over 4.1 ±2.4 yrs. For the 23 ‘progressors’ the most frequent indication for follow on CT was to investigate worsening symptomatic breathlessness (52%) and decline in lung function parameters (28%). All those who progressed to ‘definite’ and ‘probable UIP’ were diagnosed clinically, with IPF after discussion in the ILD MDT; five of which underwent surgical lung biopsy to attain definitive diagnosis. Therefore 53% (17 of 32) of our evaluable iUIP cohort (i.e. those who had follow-on CTs) or 35% (17 of 48) of all iUIP patients (if those who did not have a follow-on CT were included) progressed to a clinical diagnosis of IPF over a mean period of 3.9 ±1.9 years. These findings are summarized in Figure 1.

**Figure 1.**
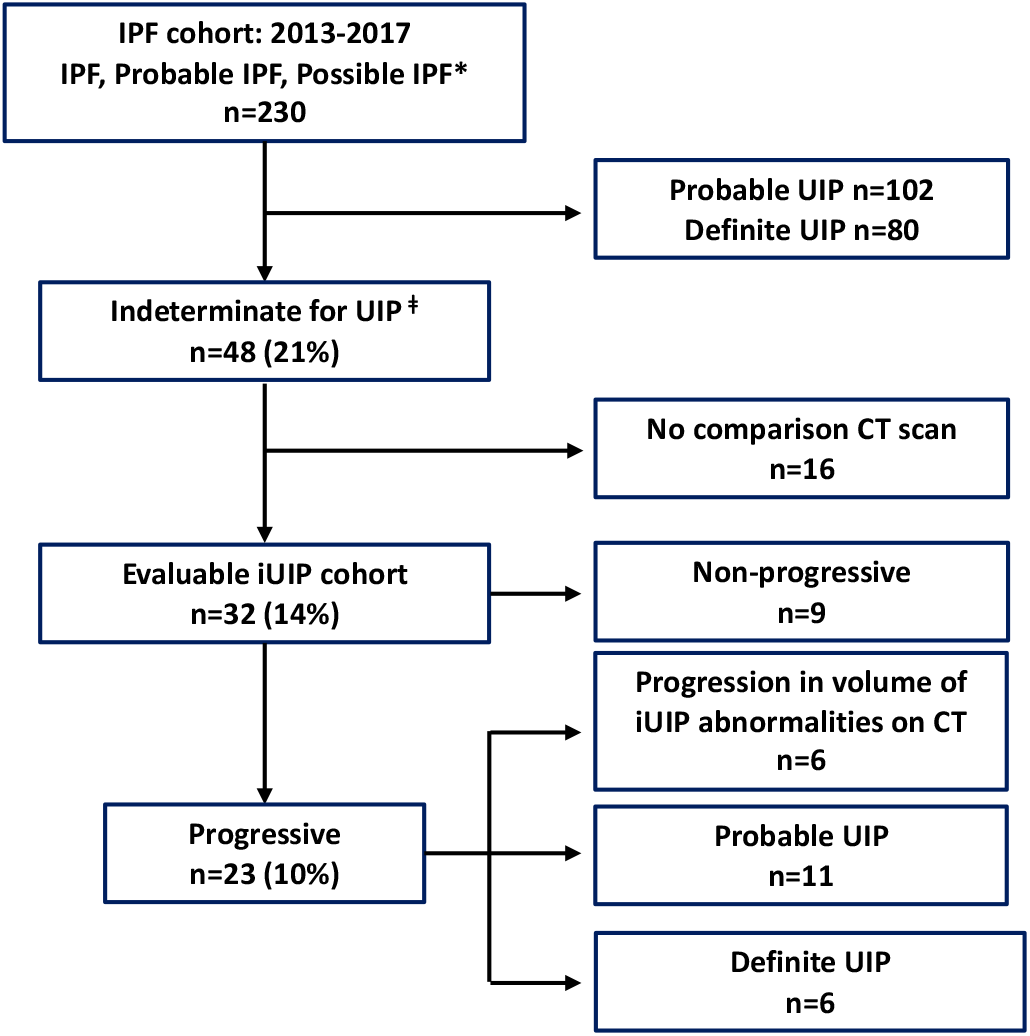
Flow diagram of radiographic progression of UIP within the IPF cohort (n=230). *; clinical diagnosis as per 2011 IPF guideline.^4^ ǂ; as per 2018 IPF guideline.^3^ UIP, Usual interstitial pneumonia; IPF, Idiopathic pulmonary fibrosis.

12 of 48 iUIP cases (25%) died during follow up. The mean time from initial CT reporting iUIP to all-cause mortality was 4.6 ±2.9 years. Respiratory-related deaths were confined to the progressive iUIP group; these accounted for 6 of the 9 deaths in this group; two to pneumonia and four to end-stage IPF. There was a trend to a greater number of hospitalisation events (39% vs 22% [OR 2.25, CI 0.40-12.32) and greater smoking history (86% vs 67% [OR 3.2, CI 0.59-15.9]) in the progressive iUIP group (Table 1). FVC and TLCO values at initial CT were not different between the progressor and non-progressor groups. However, at 1 year from initial CT, mean change in FVC for the ‘non-progressor’ group was -0.03 (±0.26) litres vs -0.26 (±0.39) litres in the ‘progressive’ group; p=0.16 (Figure 2A). Median survival for patients with an initial CT of iUIP was also significantly better than patients with a first CT demonstrating either ‘probable’ or ‘definite UIP’ (Figure 2B). Demographic data, physiological indices, and comorbidity profiles in the progressor and non-progressor iUIP groups are shown in Table 1.

**Table 1.**
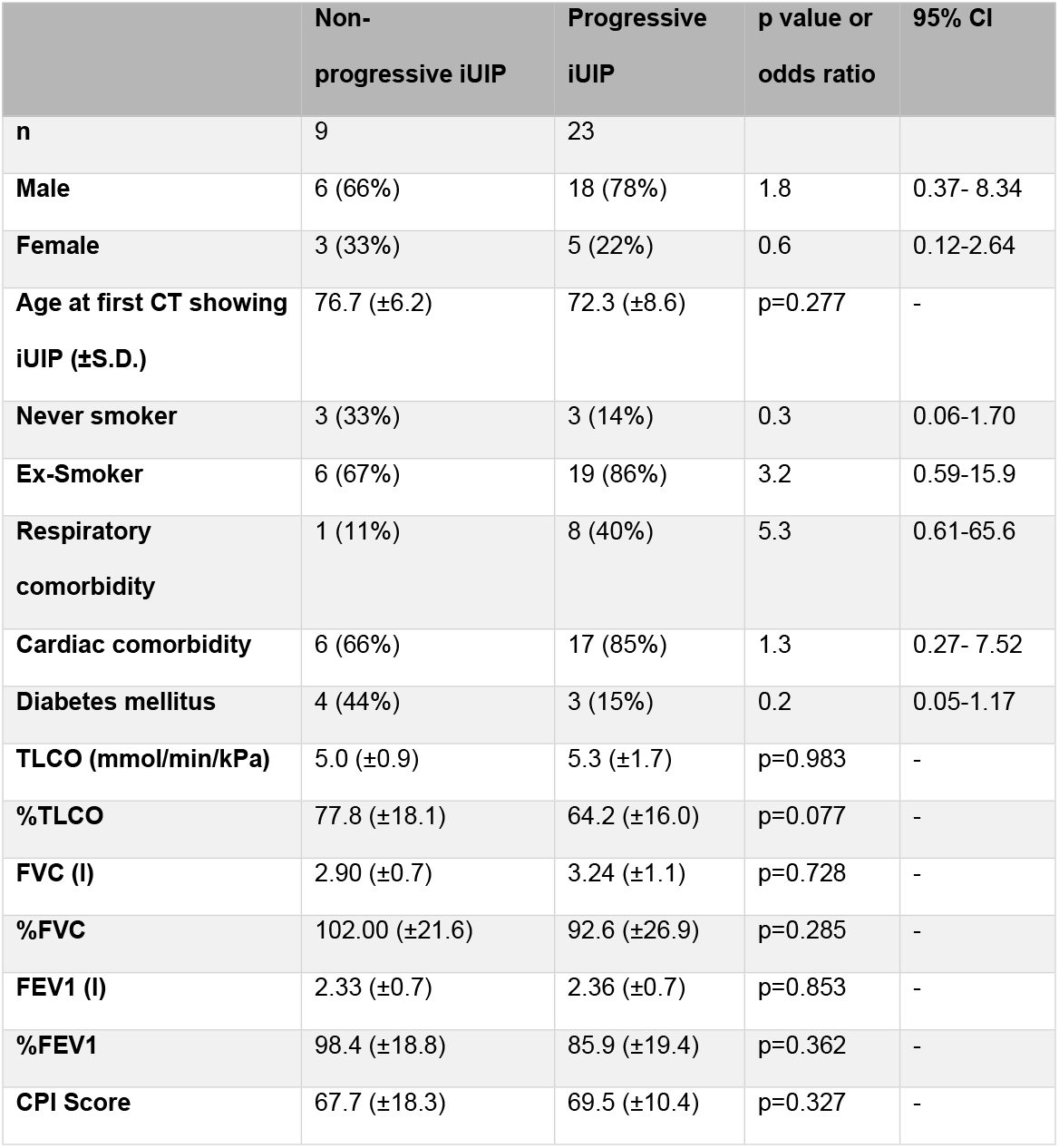
Characteristics for iUIP patients who had at least two CT scans (n=32), at the point of initial CT when iUIP was identified; divided into progressor and non-progressor groups. % in parenthesis is proportion of specified group. Statistical analysis expressed at p value or odds ratio with 95% confidence interval. Lung function parameters refer to those measured within 3 months of first CT scan. CPI; Composite Physiological Index as calculated by Wells^10^.

**Figure 2.**
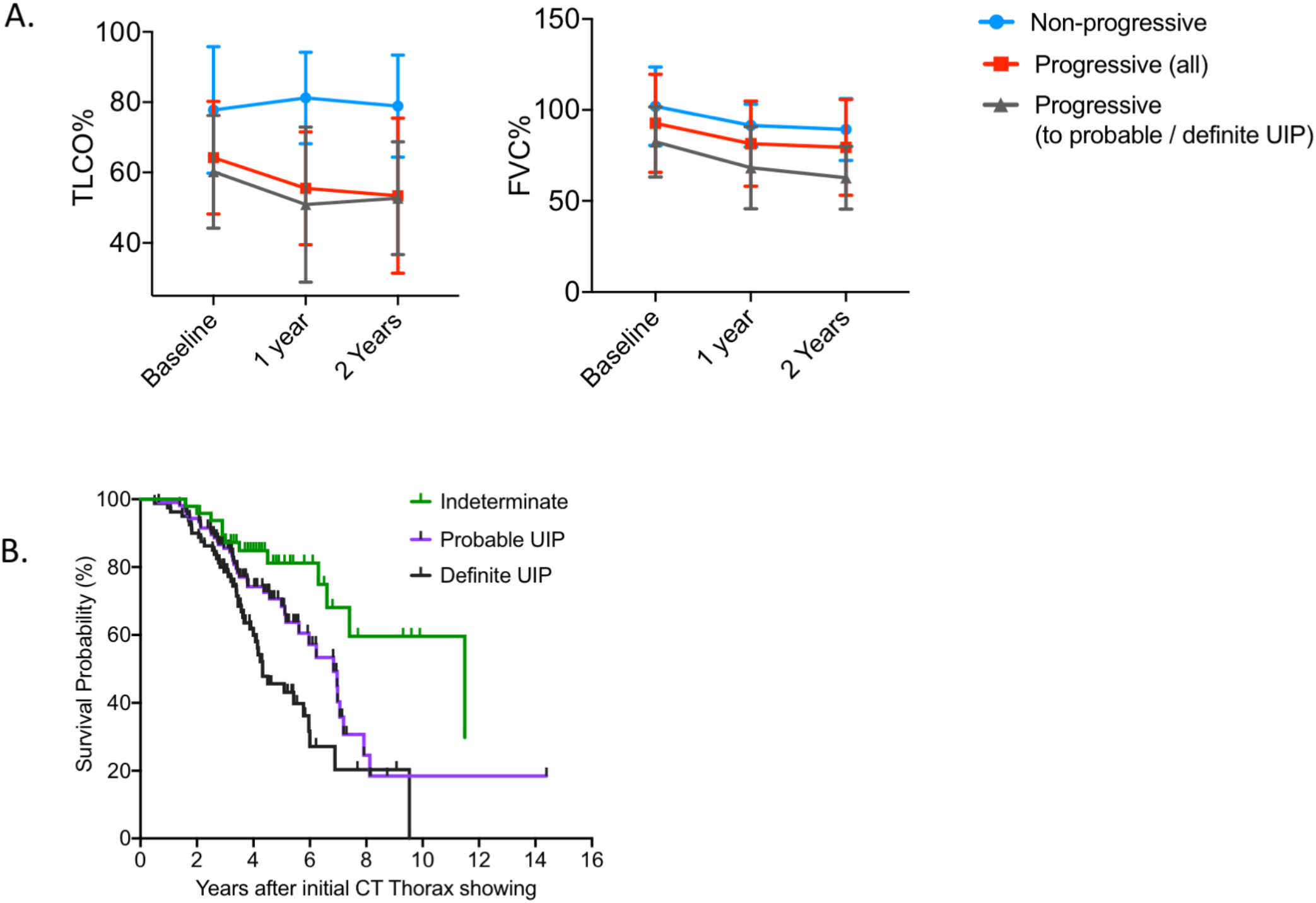
(A) Lung function progression from baseline (within 3 months of initial CT scan) for non-progressors, those who progressed in amount of disease and to ‘definite’ and ‘probable’ UIP [‘progressors (all)’], and those who progressed to definite and probable UIP only [‘progressors (to probable/definite UIP)’] Mean (S.D) values are displayed; no statistical analyses were performed.(B) Survival curve for all patients divided into those with iUIP, definite and probable UIP on thoracic CT scan at their first CT scan in the study.

### Monocyte and neutrophils but not lymphocyte levels within three months of initial CT scan are associated with progression of iUIP to definite or probable UIP CT patterns

Having characterised these subgroups, we explored potential predictor variables for progression to IPF.

A univariate analysis of the following variables [taken at the point of diagnosis (initial CT scan)] - age, gender, FVC, smoking status, monocyte count (continuous, median value or dichotomised at > and < 0.9×10^9^/l), lymphocyte count (continuous, median value, < and > 1.0×10^9^/l), neutrophil count (continuous, median value, < and > 7.5×10^9^/l) were undertaken using the Cox proportional hazard modelling method. Dichotomised values were selected from the upper limit of normal range for neutrophils and lymphocytes, and from Scott et al paper for monocytes^11^.

We determined the hazard ratios (HRs) for progression and tested their significance (using likelihood ratio test) in two settings using data from (A) all evaluable cases (n=32) and (B) only patients that progressed to IPF during the period of analysis (n=17). Apart from smoking status none of the variables violated the proportional hazards assumption of a Cox regression in setting (A). In setting (B) lymphocyte count (continuous) did; while smoking status did violate the proportional hazards assumption of a Cox regression (Table 2). Output from smoking status in setting A and lymphocytes in setting B were therefore not used.

**Table 2.**
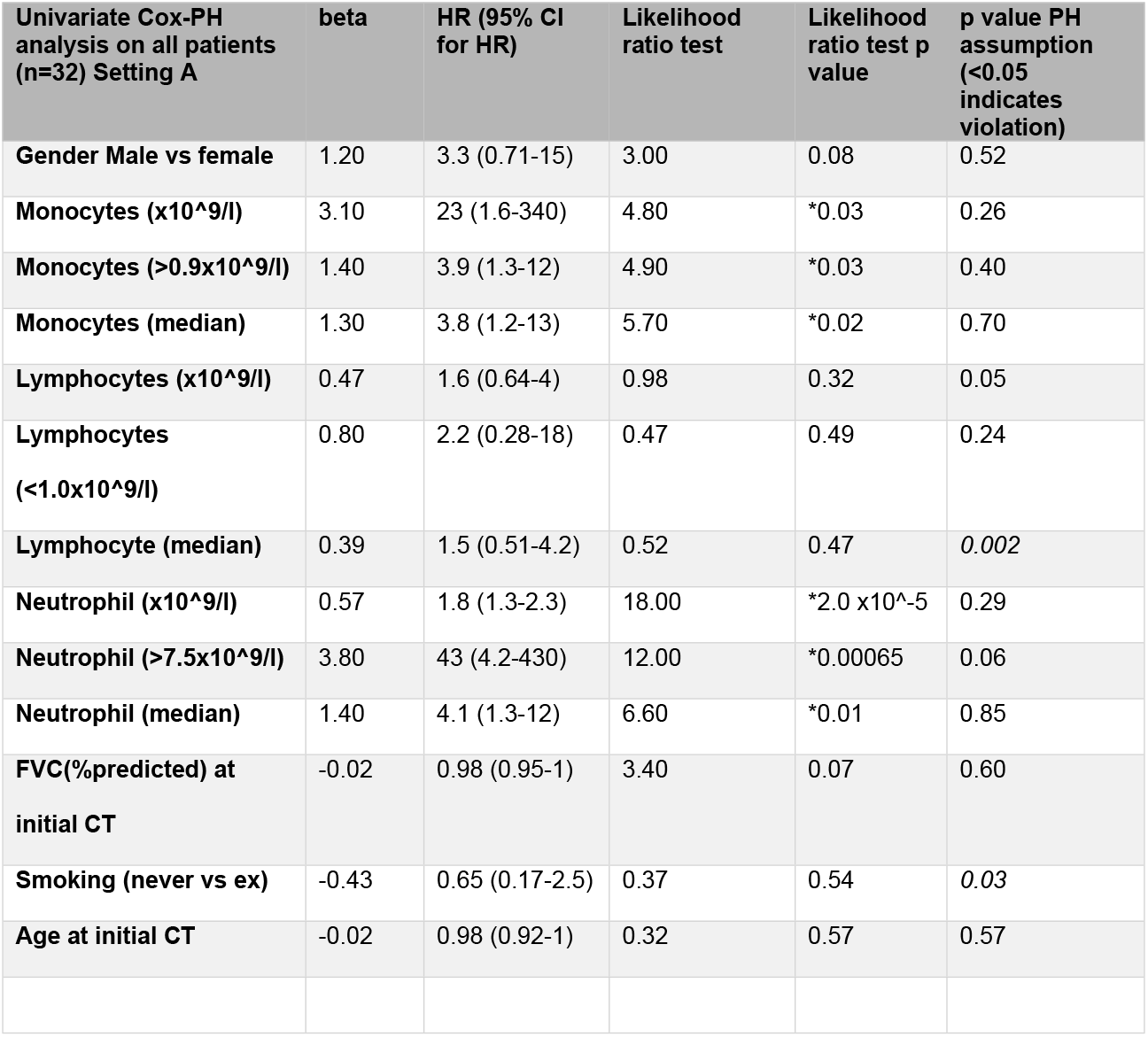
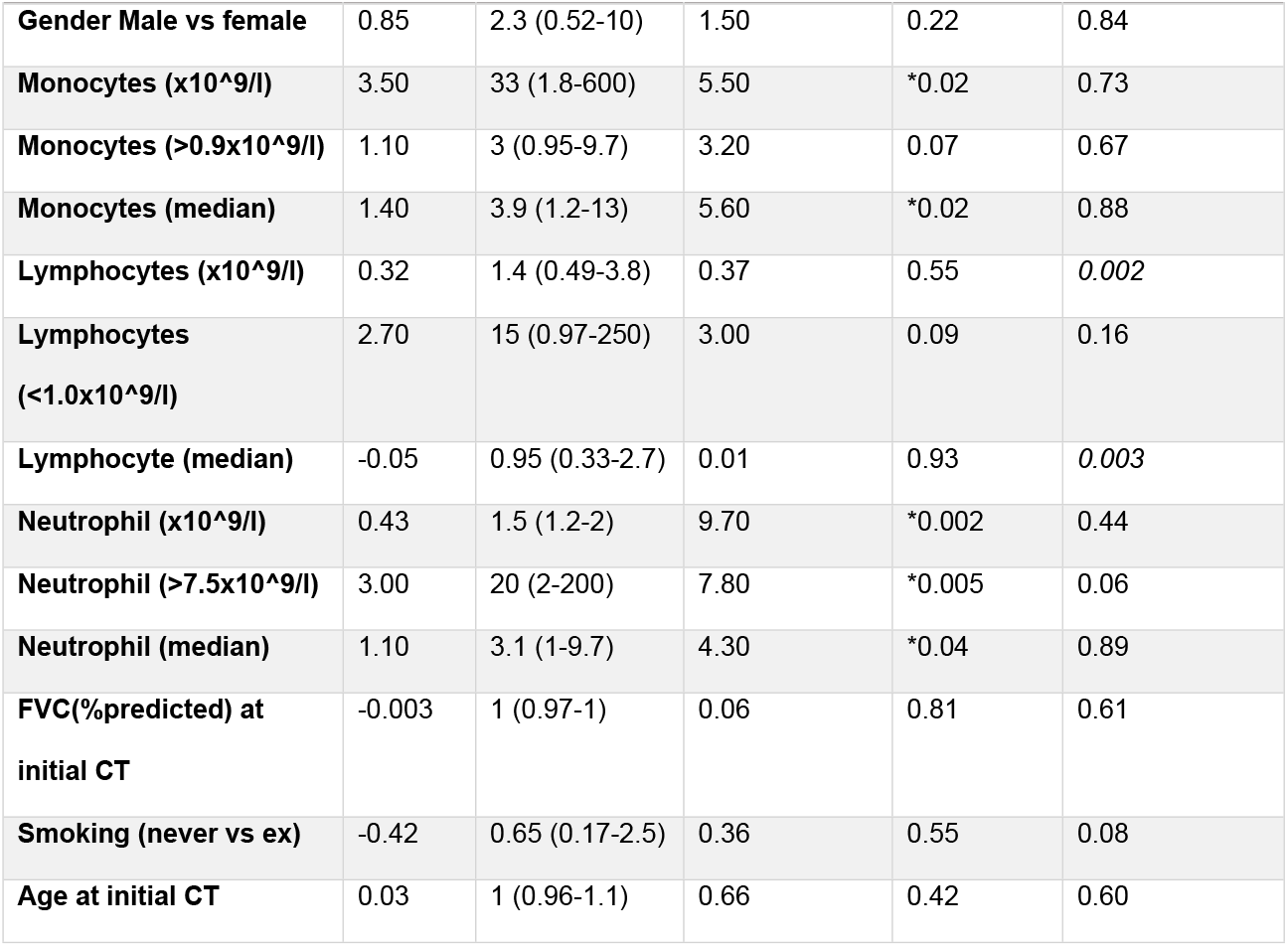
Univariate Cox Proportional hazard analysis of cohort

In both settings A and B, we found that increased neutrophils and monocytes (both binary and continuous variables) were associated with progression within the follow up period - monocytes (continuous) β=3.10; HR (95%CI) = 23 (1.6-340), p=0.03; neutrophils (continuous) β=0.57; HR (95%CI)=1.8 (1.3-2.3),p<0.001 (Table 2; and Figure 3A-B). Apart from blood leukocytes, no other variables showed significant difference. Histograms for the distribution of monocyte, neutrophil and lymphocyte values is shown in supplementary data (Figure S1).

**Figure 3.**
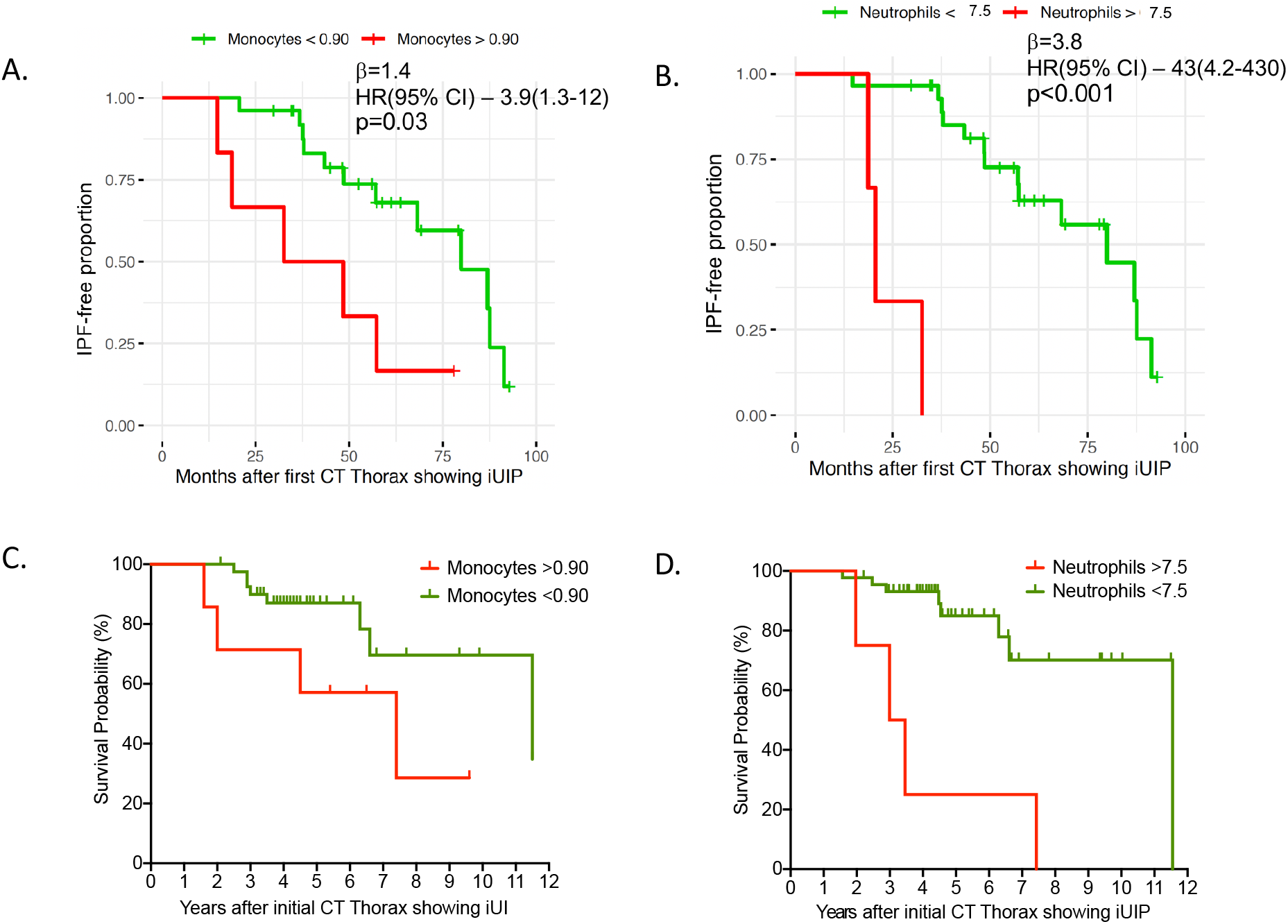
iUIP-free months in all patients (n=32) with (A) monocytes levels > and < 0.9×10^9/l and (B) neutrophils > and < 7.5×10^9/l at the point of initial CT with iUIP (univariate Cox Proportional Hazard analysis of all patients with iUIP on initial CT scan; p value analysed by Likelihood ratio test). (C) Survival curve for all patients with iUIP at initial scan (n=48) divided with (A) monocytes > and < 0.9×10^9/l and (D) neutrophils > and < 7.5×10^9/l, regardless of progression, and including those without second CT scan.

Due to the low numbers and some high monocyte, lymphocyte and neutrophil counts, we also checked the sensitivity of HR and their significance using a more balanced design by splitting the full blood cell (FBC) counts by their median values. The statistical significance and direction of effect in both settings (A) and (B) were maintained (Table 2).

We also modelled the binary monocyte, lymphocytes and neutrophil levels to account for the co-variates of gender, age and FVC in the Cox-PH multivariate mode. This multivariate analysis also preserved the significance of monocyte and neutrophil count in settings A and B (Table 3). Significance of individual covariates was reported using Wald test and overall significance of the model (compared to the alternative of no effect of any covariate) was tested using likelihood ratio test as reported by the coxph function in R.

**Table 3.**
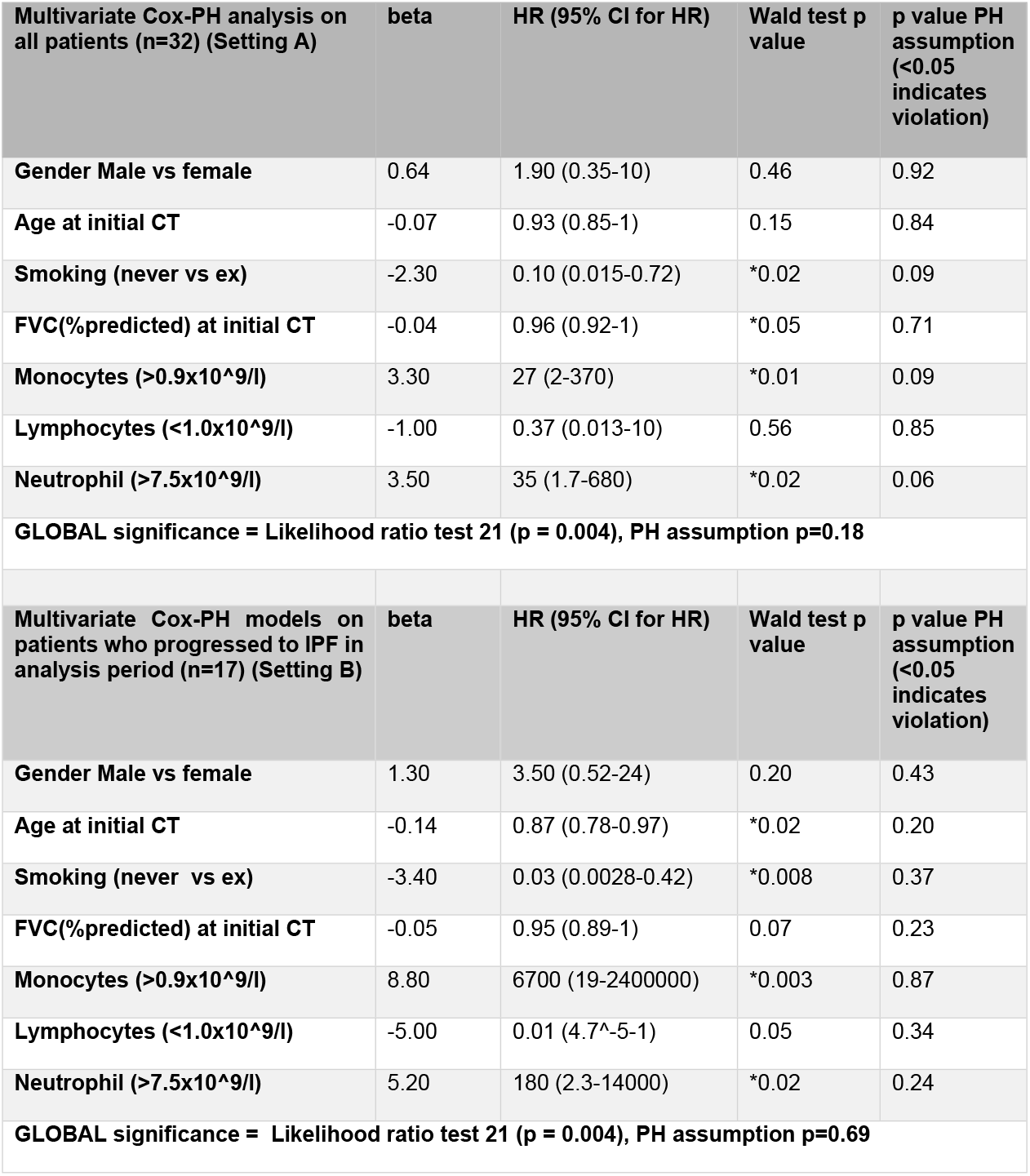
Multivariate Cox Proportional hazard analysis of cohort

The adjusted HR and confidence intervals were large but for all significant values, the lower limit range of the confidence interval was above 1, providing confidence that the HR were above 1 for monocytes and neutrophils. We do not think the absolute values of the adjusted HR and upper confidence intervals are an accurate reflection of patient risk, and are likely inflated due to the small number of patients.

Lymphocyte levels (binary or continuous) were not significantly associated with time to IPF diagnosis in any of the models.

Finally, we examined survival after categorising all cases, regardless of whether they had a follow-on CT (n=48), according to baseline monocyte count of > or <0.90 x10^9/l and neutrophils of > or < 7.5×10^9/l. We observed a trend towards shorter survival time for higher monocyte levels [HR 2.9, CI 0.59-14.51, p=0.06 Log-rank test, censoring event as death or August 2019 for survival group]; and higher neutrophils [HR 6.95, CI 0.72-66.7, p=0.0002 Log-rank test] (Figure 3C-D).

## Discussion

In this single centre retrospective analysis of iUIP progression, we observed that amongst an unselected group of 230 patients followed up for lung fibrosis, the prevalence of those with iUIP CT pattern was 21%. A minimum of 35% (if we included those who did not have a second thoracic CT scan due to lack of need), progressed to IPF within four years, with 25% death in the follow up period. These data suggest that iUIP CT pattern is an important entity, a precursor to IPF in some patients within a few years, and for these patients, there could only be a short period to intervene to prevent progression. Further analyses suggest that increased neutrophils and monocytes levels might identify this group of patients with higher risk of progression to IPF.

The prevalence of iUIP amongst patients seen in our IPF clinic is similar to the analysis from only one other study on prevalence. Diridollou et al re-categorised 89 cases with ‘possible UIP’ CT pattern and found 17% of these were iUIP^12^. In a large birth cohort study (AGES-Reykjavik, n=5320), Putman et al observed a 2.52% prevalence of iUIP patients, and demonstrated that patients with iUIP CT pattern had a greater risk of mortality compared to those without any interstitial lung abnormality (p<0.0001 [HR 1.6, 95% CI 1.3-2.0])^13^. Neither study examined how iUIP patients progress in the ensuing years from initial diagnosis.

It is noteworthy from our analysis that there is no statistical difference between the starting FVC and TLCO for the progressor and non-progressor groups of iUIP patients; though there is a trend towards lower % predicted TLCO (Table 1). The calculated CPI score which adjusts for presence of emphysema is, not unexpectedly, also similar between the two groups. However, with univariate (but not multivariate) analysis (Table 3), lower FVC at initial CT did (just) correlate with greater likelihood to progression for the individual patient, which is clinically cogent. These data suggest that factors other than severity of disease may be driving the progression of iUIP to IPF.

We chose to examine monocyte, neutrophils, and lymphocyte levels, in part, because of the potential utility in clinic due to these being routine performed blood tests but primarily because of the possible link between monocytes and mechanism of disease, as shown in Scott et al^11^ and our own work^14^. We used monocyte counts of >0.9×10^9^/L as Scott et al had identified this as the level of monocyte, above which was associated with higher mortality risk in IPF patients. In Fraser, Denney et al^14^ we observed that monocytes in IPF patients showed type 1 interferon primed phenotype which could account for more robust and potentially injurious response to the alveolar epithelium when triggered, during for example, a viral infection. This study provides impetus to investigate the possibility that neutrophils (which were not investigated in Scott or Fraser, Denny studies) could also be involved.

There are several limitations to our study. The monocyte, neutrophil and lymphocyte levels were measured at one point (nearest to the CT scan). This is in keeping with work from other much larger studies ^11^ but there is a risk that the values are not representative of the steady state values, particularly in a necessarily small cohort as ours. In further studies, it will be useful to have repeated samples over 6-12 months to determine if the neutrophil, lymphocyte and monocyte values are representative for the patient, and reduce bias towards the possibility of levels linked to an infective episode, for example.

The most obvious limitation is the small number of patients. This probably reflects the relatively small proportion of iUIP scans that are referred for clinical assessment without delay. These patients are often asymptomatic in the early stages of their interstitial lung disease.^15^ However, it is becoming increasingly recognised that a proportion of ILAs will progress to clinically significant ILD.^15^ Furthermore, identification of ILAs is predicted to increase with implementation of lung cancer screening and increased use of CT for other diagnostic purposes.^16^ This may increase reporting of iUIP in the future, making the need for biomarkers that can risk-stratify for progression even more pertinent.

In our retrospective study, not all follow on CT scans included in analysis were undertaken in a uniform timeframe across the cohort. With exception of the lung nodule surveillance imaging most serial CTs were performed according to clinical indication and where there was concern for objective deterioration. A greater proportion of repeat CT scans were performed to investigate symptomatic change in the ‘progressor’ group, and this may have introduced bias towards detection of progression. We also acknowledge that length of follow up is shorter in the non-progressor group, although not significant (p=0.06). However, it has been well documented that initial progression particularly during first 6 to 12 months is predictive of progression of fibrotic ILDs.^17,18^. Therefore a follow up for 2 years or more, as seen in the majority of our cohort, may be sufficient to identify patients with a progressive phenotype.

The small sample size limits the power of this study and has contributed to some of the large hazard ratios and confidence intervals observed. Therefore, our findings are primarily indicative rather than definitive signals. The values of the HR cannot be interpreted as an absolute numerical risk but rather an indication that the risk exist and that it is statistically significant since the lower limit of the 95% confidence interval is above 1. Consistent findings in proportional hazards modelling by both univariate and multivariate analyses also lend support to the contribution of monocytes and neutrophil levels to progression to IPF. As the numbers of patients with iUIP CT pattern disease are small, a multi-site cohort will be required to confirm these findings. Nevertheless, our findings suggest that at least some patients with iUIP CT pattern could be patients with early IPF and their disease progression could be linked to higher levels of monocytes and neutrophils. Further studies could validate the use of blood monocytes and neutrophils as biomarkers for iUIP patients who are at higher risks of progression to IPF and all-cause mortality.

## Data Availability

All relevant data is included within the manuscript and supplementary section.

## Acknowledgement

The study was funded by the National Institute for Health Research (NIHR) Oxford Biomedical Research Centre (BRC). LPH is supported in part by the MRC UK (MC_UU_00008/1). Correspondence to Prof Ling-Pei Ho.

## Authors contribution

AA conducted analysis, interpreted data and wrote the paper, AR, AP, HB, KD and RB collected data, R Benamore and RH discussed clinical diagnosis of patients, VI led statistical analysis and interpretation of data, LPH conceived project, interpreted data, wrote paper and supervised the study.

---

**Supplementary Figure S1.**
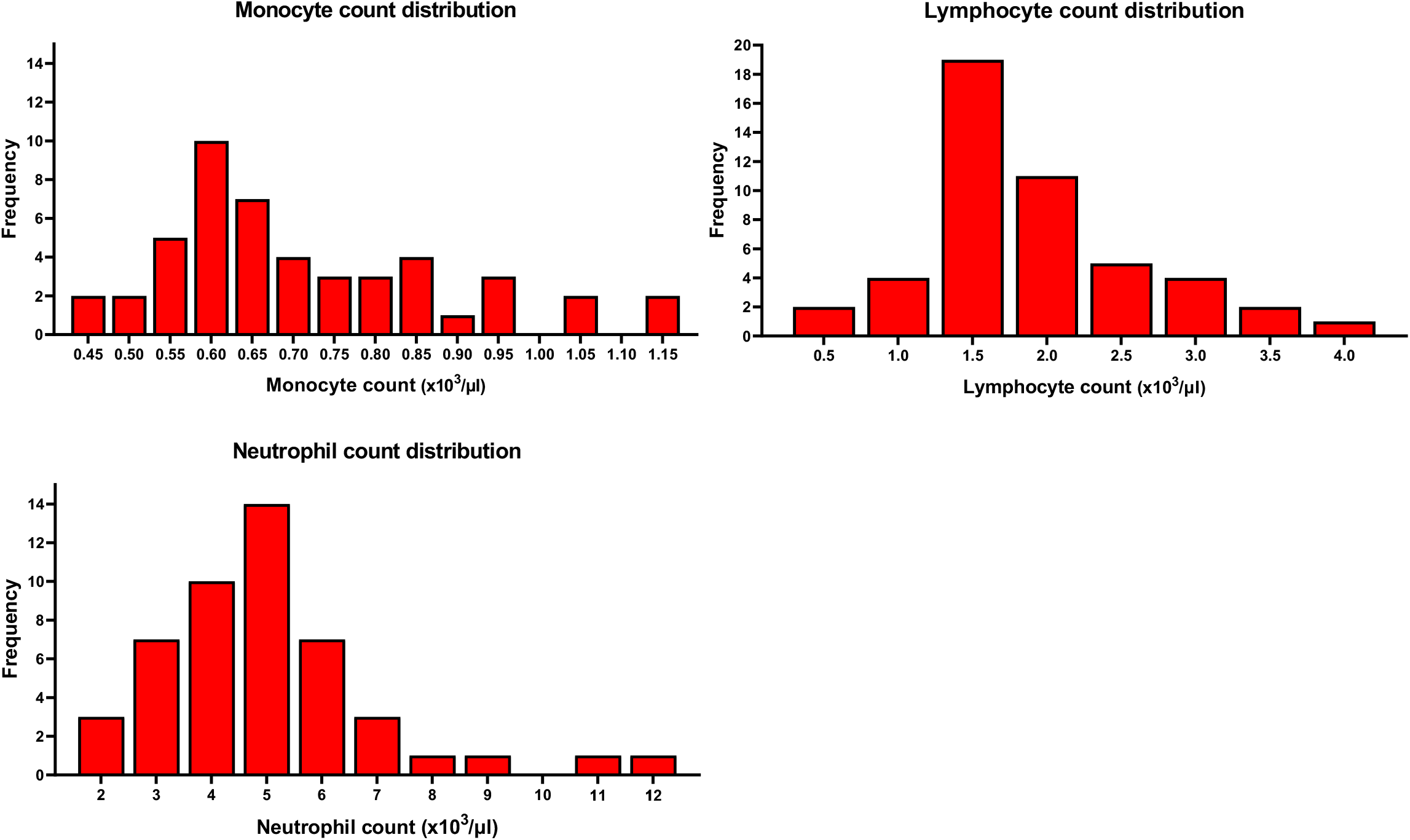
Blood leukocyte distributions

## Notes

### Competing Interest Statement

The authors have declared no competing interest.

### Author Declarations

Ethical approval The study was part of a study to examine the factors associated with disease progression in IPF (ethical approval 14/SC/1060 from the Health Research authority and South-Central National Research Ethics Service).

